# The *SORL1* p.Y1816C variant causes impaired endosomal dimerization and autosomal dominant Alzheimer’s disease

**DOI:** 10.1101/2023.07.09.23292253

**Authors:** Anne Mette G. Jensen, Jan Raska, Petr Fojtik, Giulia Monti, Melanie Lunding, Simona Vochyanova, Veronika Pospisilova, Sven J. van der Lee, Jasper Van Dongen, Liene Bossaerts, Christine Van Broeckhoven, Oriol Dols, Alberto Lléo, Luisa Benussi, Roberta Ghidoni, Marc Hulsman, Kristel Sleegers, Dasa Bohaciakova, Henne Holstege, Olav M. Andersen

**Author notes:** Corresponding author: Olav M. Andersen.

## Abstract

Truncating genetic variants of *SORL1*, encoding the endosome recycling receptor SORLA, have been accepted as causal of Alzheimer’s disease (AD). However, most genetic variants observed in *SORL1* are missense variants, for which it is complicated to determine the pathogenicity level because carriers come from pedigrees too small to be informative for penetrance estimations. Here, we describe three unrelated families in which the *SORL1* coding missense variant rs772677709, that leads to a p.Y1816C substitution, segregates with Alzheimer’s disease. Further, we investigate the effect of SORLA p.Y1816C on receptor maturation, cellular localization and trafficking in cell-based assays. Under physiological circumstances, SORLA dimerizes within the endosome, allowing retromer-dependent trafficking from the endosome to the cell surface, where the luminal part is shed into the extracellular space (sSORLA). Our results showed that the p.Y1816C mutant impairs SORLA dimerization in the endosome leading to a strong decrease in trafficking to the cell surface, resulting in decreased sSORLA shedding. Furthermore, we find that iPSC-derived neurons with engineered p.Y1816C mutation have enlarged endosomes, a defining cytopathology of AD.

Our studies provide genetic as well as functional evidence that the *SORL1* p.Y1816C variant is causal for AD. The high penetrance of the mutation suggests this mutation should be considered in clinical genetic screening of multiplex early-onset AD families.

## INTRODUCTION

Alzheimer’s disease (AD) is the world leading cause of dementia, affecting almost one-third of the population aged 85 and over ^1^. Initial genetic studies of the early-onset, familial form of the disease implicated three genes (*PSEN1* (*presenilin1*), *PSEN2* (*presenilin2*), and *APP* (*amyloid precursor protein*), encoding proteins involved in the APP processing pathway and the production of the amyloid brain deposits that are a histological hallmark of AD ^2^. More recently, disease-causing variants of these genes when modelled in iPSC-derived neurons also phenocopy the defining cytopathology of AD ^3^, which is endosome enlargements ^4, 5^.

During recent years, *SORL1* (*sortilin-related receptor-1*) has emerged as another critical gene implicated in the development of AD and, more recently, suggested as a cross-disease gene also involved in Frontotemporal Lobar Degeneration and Lewy body disease ^6^. Truncating variants of the gene have been found almost exclusively in AD patients and are now considered causal for dementia ^7, 8^. Moreover, animal and cellular models with *SORL1* deficiency replicate many of the disease hallmarks, including APP processing defects and endosome swelling ^9–12^. However, almost 80% of the rare variants observed in the *SORL1* gene are missense variants, and of all genes in the genome, missense variants in *SORL1* have the strongest association with an increased risk of AD ^8^. Each unique missense variant in *SORL1* has its individual effect on AD risk, ranging from benign to highly pathogenic, and recently a rare variant was identified in an Icelandic family that leads to autosomal dominant AD ^13^. However, for most carriers of rare missense variants a clear family history of AD is lacking, thereby making it difficult to establish the pathogenicity of *SORL1* missense variants. Therefore, different functional assays are needed to provide evidence for the pathogenic impact of the variants on protein function.

The *SORL1* gene encodes the 250 kDa, highly glycosylated type-1 transmembrane protein SORLA that is known to be involved in the retromer-dependent recycling of endosomal cargo to the cell surface, including the neuronal cargo proteins APP and GLUA1 ^14, 15^. In fact, impaired SORLA activity has been shown to lead to increased endosomal processing of APP and Aβ production ^9, 16, 17^. Furthermore, the receptor requires trafficking in the endosomal compartments for the maturation of its attached glycosylations, before it can be cleaved by TACE at the cell surface to generate the soluble sSORLA fragment ^18^. Therefore, it has been suggested to determine receptor maturation and shedding to assess if a genetic missense variant in *SORL1* is pathogenic ^19, 20^.

SORLA uses its luminal domains to bind and traffic cargo along the inside of the tubules that connect the endsosome with the cell membrane, while the C-terminal tail protrudes on the outside of the tubule membrane, where it binds the retromer core complex consisting of the trimeric VPS26-VPS35-VPS29 proteins ^21, 22^. Recently, we showed that SORLA can dimerize (and possibly form higher order oligomers), when located to endosomes, and that it is mainly when SORLA is in its dimerized (or multimeric) form that it can bind the retromer complex. Similar to other receptor proteins with a VPS10p-domain ^23^, also SORLA can make a VPS10p-domain dimer ^24^, but more importantly SORLA can also dimerize through its six Fibronectin-type III (3Fn)-domains, which correspond to the receptor part that is most proximal to the membrane ^22^ (**Fig. 1A**).

**FIGURE 1:**
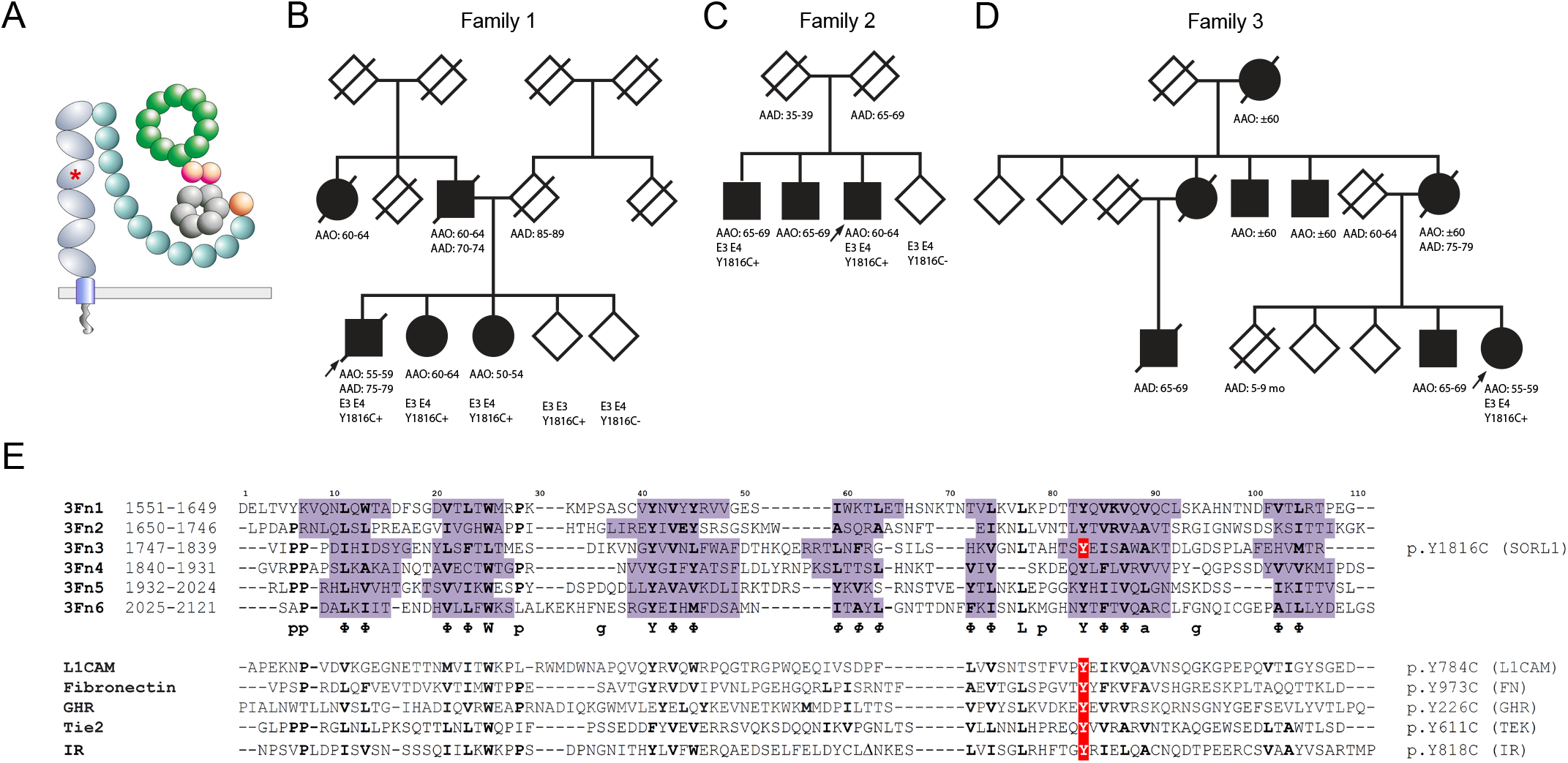
SORL1 p.Y1816C segregates with Alzheimer’s Disease in three unrelated families. (A) Schematic representation of full-length SORLA showing the domain architecture and predicted folded conformation. The location of the p.Y1816C mutation in the third 3Fn domain is indicated by a red asterisk. (B-D) Pedigrees on family 1 (B), family 2 (C) and family 3 (D) with AD and SORL1 p.Y1816C. Probands are indicated by an arrow. AAO: age at onset; AAD: age at death. Y1816C+ indicates a carrier, Y1816C-indicates a non-carrier. APOE genotype is indicated by E3 E3 or E3 E4 when determined. Vertical bar indicates affected by hearsay.(E) 3Fn-domain sequence alignment for the six SORLA domains together with homologous domains with disease-causing mutations identified at an identical domain position: L1CAM, FN, GHR, TEK, and IR. The β sign within the IR sequence represents a deletion of a domain insert for this protein.Each of these mutations lead to substitution of the conserved tyrosine at position 83 – in the so-called *tyrosine corner* – with a cysteine residue.

Here, we report a *SORL1* variant, p.Y1816C, that affects a tyrosine amino acid in the 3rd 3Fn-domain and which is highly conserved across all 3Fn-domain sequences. We observed this variant in three unrelated families (family 1, 2 and 3), and while AD affected family 3 with an inheritance pattern suggestive of autosomal dominant AD, we observed that the variant segregated with AD in thepedigrees from family 1 and 2. We found that the p.Y1816C substitution leads to a strong decrease in receptor maturation, cell surface localization, and sSORLA shedding. While we observe a small increase in ER localization of the mutant receptor, the majority of p.Y1816C protein was able to sort to the endosomes. However, once in the endosome, the mutant receptor fails to dimerize, rendering it unable to sort to the cell surface as part of the retromer-dependent recycling pathway. The failure to engage in endosomal recycling was further supported in iPSC-derived neurons since mutation of the endogenous WT *SORL1* to the p.Y1816C mutant using CRISPR/Cas9 led to endosome swelling, similar as previously observed for truncating *SORL1* variants ^10, 11^. Together, our findings suggest that SORLA must dimerize at its 3Fn-domains in order to function in retromer-dependent endosomal recycling.

## METHODS

### Ethical Oversight

The Medical Ethics Committee of the Amsterdam University Medical Center gave ethical approval for this work. All participants and/or their legal representatives provided written informed consent for participation in clinical and genetic studies.

### Molecular genetic analyses

The *SORL1* variant rs772677709 (11:121485607:A>G, GRCh37) leading to the p.Y1816C substitution in the pedigree of family 2, the proband from family 3 and the proband from family 1 was reported previously ^7, 25^. To determine segregation of *SORL1* p.Y1816C in the pedigree of family 1, exon 41 was sequenced on genomic DNA of four siblings of the proband using PCR amplification followed by Sanger sequencing using the BigDye termination cycle sequencing kit v3.1 on the ABI 3730 DNA Analyzer. Sequences were analyzed using Seqman (DNAstar, WI, USA) and NovoSNP software ^26^. In addition, APOE e2/3/4 status was determined in all sibs and the proband was screened for pathogenic mutations in APP, PSEN1 and PSEN2 using multiplex PCR amplification followed by sequencing on an Illumina MiSeq platform. For the pedigree of family 3, segregation could not be tested due to DNA unavailability.

For haplotype sharing analysis between the probands from the 3 families, seven short tandem repeat (STR) markers with heterozygosity ≥ 70% were selected in the region from chr11:117,644,118 – 126,971,788 spanning 9.3 Mb around the *SORL1* gene. STR markers were genotyped on genomic DNA of the three probands and three unrelated healthy individuals using multiplex PCR amplification with fluorescently labeled primers followed by capillary fragment analysis on an ABI 3730 DNA Analyzer (Thermo Fisher Scientific, MA, USA). Haplotype sharing analysis was further performed on single nucleotide polymorphism (SNP) genotypes in the *SORL1* gene for the probands from family 2 and 3, SNP genotypes were extracted from Illumina GSA microarray data that were previously generated ^27^.We extracted all imputed variants (imputation quality >0.3) in the region chr11:121.450.314-121.635.763 from these two individuals, where in total 184 variants were at least seen heterozygote to the reference panel, with no pattern of heterozygote varaints emerged (**Supplementary Figure S2**. For the proband from family 1, microarray SNP data were not available.

### Plasmids and constructs

The untagged WT and GFP-tagged SORLA expression constructs have been described previously ^28, 29^. SORLA Venus plasmids (pDEST-SORLA-V1 and pDEST-SORLA-V2) were a kind gift from Johanna Ivaska, University of Turku and specified previously ^30^. The Venus-tagged SORLA 3Fn-minireceptor constructs (3Fn-mini-V1 and 3Fn-mini-V2), pRP(Exp)-CMV_T7>3Fn-Mini:Linker:V1 and pRP(Exp)-CMV_T7>3Fn-Mini:Linker:V2, were constructed by VectorBuilder Inc. The vector IDs are VB221204-1293tpe and VB221204-1288bvv respectively that can be used to obtain detailed information about these vectors.

### Site-directed Mutagenesis

The SORLA expression constructs containing the p.Y1816C mutation was generated by Quickchange XL site-directed mutagenesis (#20052, Agilent) with SORLA WT, SORLA GFP, pDEST-SORLA-V1, pDEST-SORLA-V2, 3Fn-mini-V1 or 3Fn-mini-V2 as templates and amplified using the primer pair (fwd: 5’-GGCAATCTGACAGCTCATACATCC**TGT**GAGATTTCTGCCTGGGCCAAGACTG-3’, rev: 5’-CAGTCTTGGCCCAGGCAGAAATCTC**ACA**GGATGTATGAGCTGTCAGATTGCC-3’). All steps were performed following the protocol from the manufacturer.

### Cell culture

HEK293 cells (ATCC) were cultured in Dulbecco’s Modified Eagles Medium (Gibco) supplemented with 10% fetal bovine serum (FBS) and 1% penicillin/streptomycin in a humidified 5% CO_2_ incubator at 37℃. Transient transfections were performed using Fugene 6 Transfection Reagent kit (Promega) with a Fugene:DNA ratio of 3:1.

For SORLA maturation and shedding: HEK293 cells were seeded at a density of approximately 400.000 cells per well of 6-well plates 24 hours before transfection with 3 µg of plasmids encoding SORLA WT or SORLA p.Y1816C. Subsequently, the medium was changed to serum-free medium 24 hours post transfection and cultured for another 48 hours before harvesting conditioned medium and cell lysates.

### Immunoblotting

HEK293 cells were lysed on ice for 1 hour in lysis buffer (1M Tris HCl pH 8.1, 0.5M EDTA, 1% Triton-X-100, 1% NP-40) containing protease inhibitor (Complete^TM^, Roche). Total protein concentration in lysates was determined using the bicinchoninic acid assay kit (Sigma). Protein samples were mixed with SDS sample buffer supplemented with 5% β-mercaptoethanol followed by incubation at 95℃ for 5 min. SDS-PAGE separation of equal amounts of protein was performed using precast 4-12% Bis-Tris gradient gels (Thermofisher) at 84V and 100W for 10 min followed by 160V and 100W for 1 hour and 15 min. Subsequently, proteins were transferred to nitrocellulose membranes with iBlot^TM^ 2 Gel Transfer (Thermofisher) and incubated in blocking buffer (Tris-Base 0.25M, NaCl 2.5M, skimmed milk 2%, Tween-20 2%) for 1 hour at room temperature. The membranes were incubated in primary antibodies overnight at 4℃ followed by incubation in HRP-conjugated secondary antibodies (DAKO) diluted 1:1500 for 1 hour at room temperature the day after. Proteins were visualized by ECL prime detection reagent (Sigma) using the iBright^TM^ imaging system (Thermofisher). Antibodies: anti-LR11 (#612633, BD Transduction Laboratories) and anti-β-actin (#A5441, Sigma).

For iPSCs protein lysis and Western blot were performed as described previously ^31^. Briefly, protein samples were lysed in 1% SDS-lysis buffer, their concentration was measured with DC™ Protein Assay (Bio-Rad), mixed with 10x Laemmli buffer, and incubated at 95 °C for 10 min. Proteins were separated on 10% Acrylamide gel and transferred onto PVDF membranes (Merck). Membranes were blocked and incubated with antibodies in 5% skimmed milk in TBS with 0.1% Tween 20. Antibodies are listed in **Supplementary Table S1**. The results were visualized via ECL™ Prime (Amersham) using ChemiDoc™ Touch Imaging System (Bio-Rad). The density of bands was quantified using ImageJ software ^32^.

### Immunocytochemistry (ICC)

HEK293 cells were seeded on poly-L-lysine-coated (0.1%, Sigma) glass coverslips at a density of approximately 40.000 cells per well of a 24-well plate. After 24 hours cells were transfected with 500 ng of the indicated SORLA expression constructs and cultured for another 24 hours. Then, cells were fixed in 4% paraformaldehyde (PFA) for 10 min at room temperature followed by washing in PBS pH 7.4. Subsequently, cells were washed 2x in PBS + 0.1% Triton-x-100 (PBST) and incubated in blocking buffer containing 10% FBS for 30 min at room temperature. Next, cells were incubated in primary antibodies diluted in blocking buffer overnight at 4℃ then washed 2x in PBST pH 7.4. Cells were then incubated in Alexa Fluor conjugated secondary antibodies (Invitrogen) diluted 1:500 in blocking buffer for 1 hour at room temperature followed by sequential washes in PBST pH 7.4, PBS pH 7.4 and ddH_2_O for 10 min each. Finally, the cells were mounted on glass slides with mounting medium containing DAPI (Sigma) for counterstaining of nuclei and imaged using a Zeiss LSM800 confocal microscope. Antibodies: polyclonal rabbit anti sol-SORLA at 1:300 (in-house, pAb 5387, Aarhus University), anti calnexin at 1:100 (#BD610523, BD Biosciences), anti early endosome antigen 1 (EEA-1) at 1:100 (#BD610456, BD Biosciences), and anti vacuolar protein sorting 35 (VPS35) at 1:200 (EB06268, Everest Biotech).

iPSC-derived neurons were plated and cultured on glass coverslips (ThermoFisher) coated with Matrigel® hESCs-qualified matrix (Corning). Cells were then fixed in 4% paraformaldehyde and permeabilized by 0.1% Triton-X100 followed by the incubation in the blocking buffer (1% BSA, 0.03% Tween 20 in PBS). Incubation with primary antibodies was performed at 4 °C overnight, and secondary antibodies were incubated with cells at room temperature for 1 h. Primary and secondary antibodies (**Supplementary Table S1**) were diluted in blocking solution. Nuclei were counterstained with DAPI (Sigma), and Mowiol (Sigma) was used as a mounting media.

### BiFC

HEK293 cells were seeded on glass coverslips coated with poly-L-lysine 0.1% (Sigma) at a density of approximately 40.000 cells per well in a 24-well plate. The cells were cultured for 24 hours before transfection with 500 ng of either the individual SORLA Venus constructs or both simultaneously. Transfected cells were cultured for another 24 hours followed by fixation in 4% PFA for 10 min at room temperature then washed in PBS pH 7.4. Subsequently, cells were stained for VPS35 and calnexin in addition to counter-staining with DAPI as described for ICC above. Finally, the cells were visualized on a Zeiss LSM800 Airyscan, Laser Scanning Confocal microscope.

### Confocal microscopy and image processing

All images of stained HEK293 cells were acquired using a Zeiss LSM800 Airyscan, Laser Scanning Confocal microscope with 63x/1.4 plan apochromat oil immersion objective with sensitive GaAsP detection. Pinhole size was set to 1AU. Image analysis was performed using ImageJ software (Schindelin 2012). Colocalization was quantified using the JaCOP plugin (Bolte and Cordelieres 2006) in ImageJ by applying the same filtering and thresholding settings to all images and presented as Mander’s correlation coefficient (Manders 1993). Profile intensity plots were generated from representative images using ImageJ software by drawing a line spanning selected VPS35/Calnexin- and- Venus-positive structures. Then, signal intensity was plotted against distance (µm) over the line drawn and after scale calibration.

iPSC-derived neurons were imaged on a Zeiss LSM 800 Laser scanning confocal microscope, equipped with a 561 laser, using a Plan-Apochromat 63x / 1.40 OIL lens (Zeiss). Pinhole size was set to 1AU. 1437 x 1437 pixel images with 0,071 µm pixel size were acquired using GaAsp PMT detector. Alexa Fluor 594 acquisition parameters were: 565-617nm (emission wavelength range), 1.47µs (dwell time), and 700V (gain). A line average of 2 was applied. For z-stack imaging, slices were acquired with a 0.5 um z-step size.

Acquired images were analyzed using the IMARIS commercial software (v. 9.8, Bitplane, Oxford Instruments). We used the Imaris Surfaces function to evaluate the endosome size. The volume was selected as the monitored parameter, and its threshold was set manually to control. Set parameters were used for batch analysis of samples.

### GFP-trap precipitation

SORLA dimers were immunoprecipitated using GFP-Trap Magnetic Particles (#M-270, Proteintech). Transfected HEK293 cells were harvested as described for immunoblot. Total lysate from transfected HEK293 cells was incubated with 25 µl magnetic beads and incubated for 1 hour at 4℃ with end-over-end rotation. The beads were then washed 5x in wash buffer (10 mM Tris/HCl pH 7.5, 150 mM NaCl, 0.05% NP-40, 0.5 mM EDTA) and eluted in SDS sample buffer supplemented with β-mercaptoethanol before proceeding with immunoblot.

### Flow Cytometry

HEK293 cells were seeded in poly-L-lysine coated (0.1%, Sigma) wells of 6-well plates and cultured for 24 hours before transfection with 3µg of SORLA-GFP expression constructs. After another 24 hours cells were collected by trypsinization for 5 min at 37℃ and incubated in blocking buffer (PBS pH 7.4, 0.1 % BSA) for 15 min at 4℃ with end-over-end rotation. Subsequently, the cells were incubated with polyclonal rabbit anti sol-SORLA antibody (in-house, pAb 5387, Aarhus University) diluted 1:500 in blocking buffer for 1 hour with end-over-end rotation at 4℃. The cells were then washed 2x in PBS pH 7.4 and incubated in rabbit Alexa-Fluor-647-conjugated secondary antibody (Invitrogen) diluted 1:500 in blocking buffer for 30 min at room temperature with end-over-end rotation. Finally, the cells were washed 2x in PBS pH 7.4 and resuspended in FACS buffer (PBS pH 7.4, 2% FBS, 1% Glucose) before filtering through a cell strainer. Propidium iodide for detection of dead cells was added to the cells immediately before analysis. The cells were analyzed using a Novocyte 3000 flow cytometer (Agilent, Santa Clara, CA) and initial gating for live cells was performed using NovoExpress software (v. 1.5.6., Agilent, Santa Clara, CA). FlowJo^TM^ software (v. 10.8.1, BD Life Sciences) was used for subsequent analysis of flow cytometry results. The full set of gating strategies used for the flow cytometry experiments are outlined in **Supplementary Fig. S1**.

### i3N stem cell line cultivation and neuronal induction

Human induced pluripotent stem cell line i3N (i3N-iPSCs) with stably integrated doxycycline-inducible NGN2 (Fernandopulle et al., 2018) was propagated as a feeder-free monolayer culture on Matrigel® hESCs-qualified matrix (Corning) in mTeSR™1 medium (STEMCELL Technologies). The previously published protocol (Fernandopulle et al., 2018) has been used for the differentiation of i3Ns iPSCs into neurons. Briefly, (D0) cells were plated as a single cell monolayer on Matrigel® coated dish at the density of 1,5-2x10^5^ cells/cm^2^ into Induction medium (IM; **Supplementary Table S1**) supplemented with 2 µg/mL doxycycline (Sigma) and 10 µM Y-27632 ROCK inhibitor (Selleckchem). For 2 days (D2-3), IM with 2 µg/mL doxycycline was changed daily. At D3, cells were re-plated onto Poly-L-Ornithine (Sigma) coated dish with Cortical Neuron medium (CNM; **Supplementary Table S1**) at the density of 1,5-2x10^5^ cells/cm^2^. Half of the medium was changed twice a week until the sample collection at D17.

### CRISPR knock-out of SORL1 and CRISPR knock-in of SORLA p.Y1816C mutation

CRISPR-KO was performed in i3N-iPSCs by RNP complex composed of Cas9-GFP (Sigma) and previously published gRNA targeting exon 6 of the *SORL1* gene ^10^. CRISPR-KI was performed in iPSCs by RNP complex composed of Cas9-GFP (Sigma) and in-house designed gRNA and ssODN as a template for homologous recombination. We have designed these vectors with the following properties: i) Cleavage site for the restriction enzyme BtsCI (Recognition sequence GGATG), which is present in the WT *SORL1* sequence, will disappear after the insertion of p.Y1816C mutation and the insertion of silent PAM mutation; and ii) New cleavage site for the restriction enzyme AciI (Recognition sequence CCGC) will be generated in the *SORL1* p.Y1816C mutant. This approach allowed us to efficiently screen for mutant KI clones using RFLP-PCR. RNP complexes were electroporated into cells by Neon™ Transfection System (Thermo), and transfected cells were sorted for GFP signal to improve CRISPR efficiency. For the SORLA-KO, cells were sorted to 96 well-plate as single cells, and single-cell clones were then subsequently tested to express SORLA protein by Western blotting. *SORL1*-KO clones were verified by sanger sequencing (Seqme). For the SORLA p.Y1816C KI, cells were sorted to 96 well-plate as single cells, and RFLP-PCR was then performed to select clones with incorporated mutation. PCR product with WT sequence was cleaved by BtsCI (New England Biolabs), while PCR product with introduced p.Y1816C mutation was cleaved by AciI (New England Biolabs). The selected clone with successfully introduced p.Y1816C mutation was verified by sanger sequencing. Sequences of gRNAs, ssODN, and PCR primers are listed in **Supplementary Table S1**.

### RNA isolation, cDNA synthesis, and qPCR

Total RNA from cells was isolated by RNA Blue reagent (Top-Bio) according to the manufacturer’s instructions. NanoDrop 1000 (ThermoFisher) was used to determine RNA concentration and purity. Reverse transcription was performed using 0.5-1 ug of total RNA and Transcriptor First Strand cDNA Synthesis Kit (Roche) according to the manufacturer’s instructions. qPCR was performed from the cDNA samples using LightCycler® 480 SYBR Green I Master kit (Roche) in 10uL reaction on LightCycler 480 II (Roche) following optimized protocol: preincubation (95°C for 5 min), followed by 45 cycles of amplification and detection (95°C for 10 s, 60°C for 10 s and 72°C for 10 s). Ct values were calculated using the automated Second Derivative Maximum Method in LC480 software (Roche). Data were processed by calculating ΔCt (Ct_gene_ – Ct_housekeeping_) and subsequently 2^-ΔCt^. Primers are listed in **Supplementary Table S1**.

### Statistical analyses

For statistical analysis, two-tailed Student’s t-test was used and Graphpad Prism (v. 9.5.0, La Jolla, CA, USA) was used to generate graphs and to calculate p-values. P-values above 0.05 were considered non-significant (ns), while *p* < 0.05 (*), *p* < 0.01 (**), *p* < 0.001 (***), and *p* < 0.0001 (****) were considered significantly different.

All data are expressed as the mean ± standard error of the mean.

## RESULTS

SORLA is a large mosaic protein that, among other domains, consists of a cassette of six 3Fn domains where the tyrosine residue at position 1816 is located within the third 3Fn domain (**Fig. 1A**). The *SORL1* variant rs772677709 (leading to the p.Y1816C mutation) was carried by three patients diagnosed with early-onset AD and a positive family history of AD, with age at onset ranging between 50 and 69 years (**Fig. 1B-D**). SNP and STR analysis of the locus provided no evidence that probands were related, suggesting that these families may have been affected by mutations that arose independently (**Supplementary Table S2**).

The carrier from family 1 had an onset age between 55 and 59 years5, and carried the *APOE*3/4 genotype. The carrier died at an age between 75 and 79 years due to advanced dementia. No pathogenic mutations were identified in *APP*, *PSEN1* or *PSEN2*. Family history was indicative of familial clustering of AD with a pattern compatible with autosomal dominant inheritance. A diagnosis of AD was reported for one of the proband’s parents (onset age between 65 and 69 years), the affected parent’s sibling (onset age between 60 years and 64 years), and two of the proband’s siblings (onset at ages between 50 and 54 years and 60 and 64 years respectively, both *APOE*3/4). DNA was available for four siblings, revealing the *SORL1* p.Y1816C mutation in both affected siblings, as well as one unaffected sibling aged in the range of 55 and 59 years at last follow-up, this sibling was *APOE*3/3. The unaffected parent died at an age between 85 and 89 years without symptoms of cognitive decline (**Fig. 1B**).

The carrier from family 2 had AD with onset at an age between 60 and 64 years and carried the *APOE*3/4 genotype. Two affected siblings had both onset of AD at ages between 65 and 69 years. One of the children of the proband was reported by a family member to be affected by Parkinson Disease with onset at an age between 55 and 59 years (**Fig. 1C**). The parents died at ages between 35 and 39 years and 65 and 69 years respectively, without a diagnosis of dementia. We previously demonstrated the presence of the variant in an affected relative while absent from an unaffected relative ^7^.

The carrier from family 3 had an age at onset between 55 and 59 years, carried the *APOE*3/4 genotype, and was diagnosed with AD at an age between 60 and 64 years, which was confirmed by CSF biomarkers. Family history was indicative of familial clustering: a sibling of the proband developed AD at an age between 65 and 69 years, a parent developed first symptoms of dementia at an age between 60 and 64 years (died at an age between 75 and 79 years) and a grandparent was reported to have dementia at around 60 years. Three out of five of the parent’s siblings were diagnosed with early-onset dementia. A cousin of the proband died with AD at an age between 65 and 69 years. (**Fig. 1D**). No additional family members could be included for DNA-testing. No pathogenic mutations in *APP*, *PSEN1* or *PSEN2* were identified in the proband.

A recent large exome sequencing case-control study identified the p.Y1816C mutation in a total of 6 individuals, which included the proband from family 3, four individuals from ADES-FRANCE with ages at onset ranging between 44 and 75 years, and an individual in the ADSP dataset with an age at onset between 70 and 74 years. Furthermore, the GnomAD database (v.2.1.1) does not report the rs772677709 variant in the non-neuro dataset, consisting of 114,704 individuals that were *not* collected as part of a neurologic or psychiatric case/control study. When including individuals with neurological diseases, the GnomAD database reports two carriers in 125,121 individuals: a (non-Finnish) European male and in an Ashkenazi Jewish male.

At the protein level, variant rs772677709 results in a mutant SORLA protein where tyrosine-1816 is substituted with a cysteine; p.Y1816C with REVEL score of 0.897 and a CADD score of 28.7. The tyrosine is located within the third of six 3Fn-domains that are part of the SORLA luminal fragment, and its position in the domain sequence corresponds to a highly conserved residue that is required for establishing a ‘*tyrosine-corner*’ in 3Fn-domains ^33^. Furthermore, an alignment of 3Fn-domain sequences carrying pathogenic mutations identified how substitutions of the conserved tyrosine with cysteines also in homologous domains of L1CAM (p.Y784C), Fibronectin (p.Y973C), Growth-hormone receptor (p.Y226C), Tie2 (p.Y611C), and the insulin receptor (p.Y818C) can act as disease-causing (**Fig. 1E**) (summarized in our *SORL1* compendium ^34^).

Altogether, this suggest that the rs772677709 *SORL1* variant (p.Y1816C) is a pathogenic and causal variant for AD in carriers of the variant in the three families.

To investigate the functional defects associated with the SORLA p.Y1816C mutant, we performed functional experiments in cellular models. First, we used transfected HEK293 cells to determine how the mutation affects the maturation and shedding of SORLA (**Fig. 2A**). We found a significant reduction in the maturation of the mutant compared to WT receptor (mature/immature SORLA relative to WT (%): 100 ± 0 (WT) vs 65.8 ± 5.4 (p.Y1816C); p = 0.0011) as observed by a decrease in the level of the high-molecular SORLA glycosylation form (**Fig. 2B**). This was accompanied by a strong reduction in the amount of shed sSORLA in the medium (sSORLA levels relative to WT (%): 100 ± 0 (WT) vs 29.4 ± 14.6 (p.Y1816C); p = 0.0023) (**Fig. 2C**). Additionally, we had access to CSF from one index patient of the Dutch family. WB analysis of the CSF showed a low level of sSORLA when compared to CSF from an unrelated AD patient with same age of onset, gender and *APOE* genotype, but WT for *SORL1* (**Fig. 2D**). These findings agree well with a previous report suggesting that maturation of the p.Y1816C mutant protein could be impaired ^20^, as well as our recent demonstration that the level of sSORLA for this mutated protein was decreased for transfected N2a cells ^19^. The decreased level of sSORLA in the medium suggests that the mutant receptor might not be able to reach the cell surface, where SORLA is cleaved by TACE and sSORLA is produced ^18^.

**FIGURE 2:**
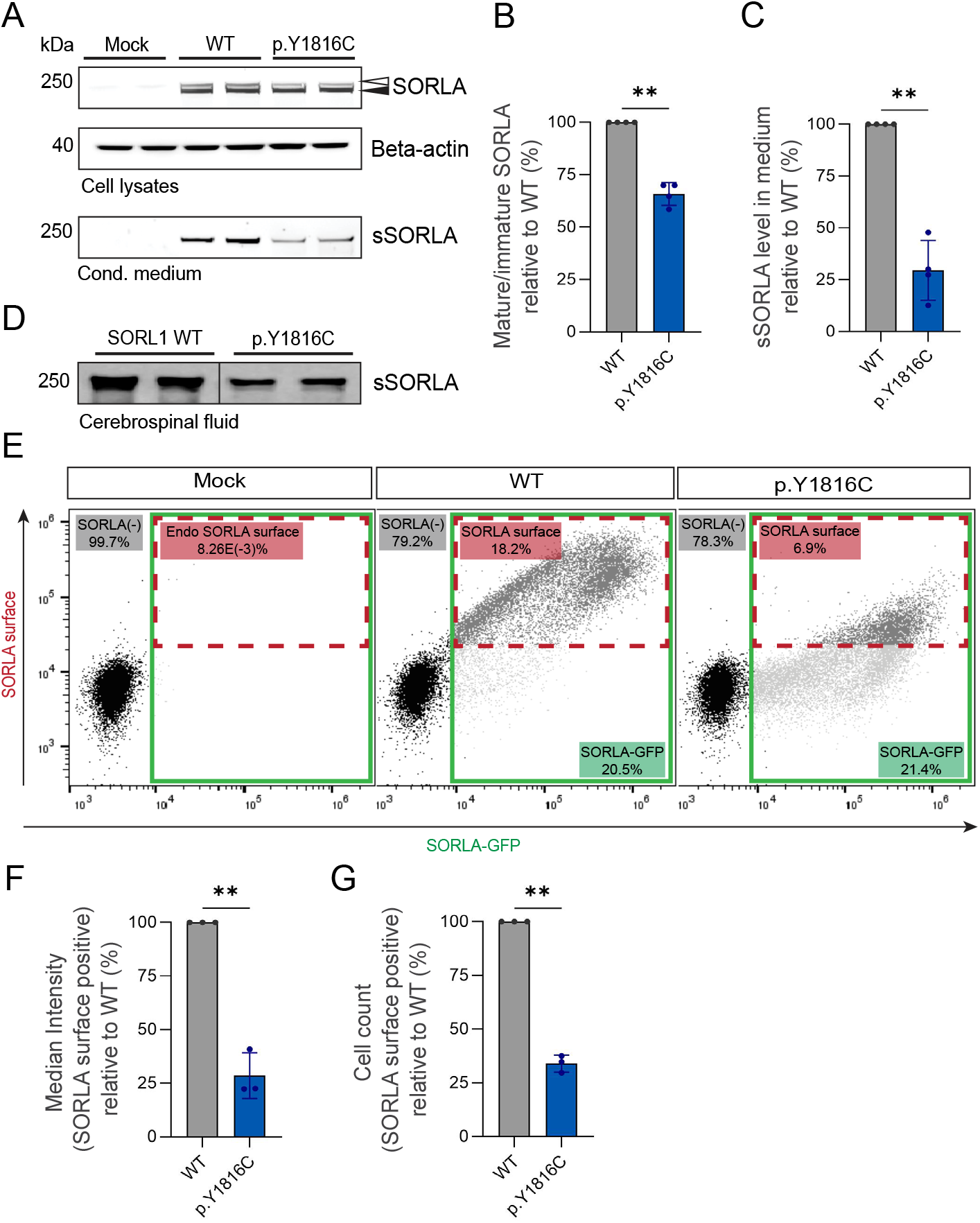
SORLA maturation and shedding is decreased for p.Y1816C variant in HEK293 cells. (A) Representative immunoblots of lysates and conditioned medium from mock transfected HEK293 cells or HEK293 cells transfected with WT or p.Y1816C SORLA constructs using antibodies against SORLA (LR11) and beta-actin. Beta-actin levels are used as loading controls (n = 8 from four independent experiments). Quantification of the SORLA maturation ratio (mature/immature) in lysates (B) and sSORLA levels in conditioned medium (C) relative to WT SORLA by densiometric scanning of immunoblots. (D) Representative immunoblots of CSF from two AD patients confirmed to be either WT for *SORL1* or to have the *SORL1* p.Y1816C mutation.(E) Representative flow-cytometry plots of either mock-transfected HEK293 cells or HEK293 cells transfected with GFP-tagged WT or p.Y1816C SORLA expression constructs. Plots show SORLA-GFP expression versus SORLA surface expression on live HEK293 cells. Green boxes represent SORLA-GFP positive cells and dashed red boxes represents cells positive for SORLA surface expression within the SORLA-GFP positive population. Numbers indicate frequencies of the different cell populations. Endo SORLA surface: endogeneous SORLA on cell surface. (n = 6 from three independent experiments). Bar graphs representing median fluorescence intensity of SORLA surface signal (F) and cell count (G) for cells within the SORLA surface positive population relative to WT. Error bars indicate mean with SD. All data were analyzed by parametric statistical tests. *p < 0.05, **p < 0.01, ***p < 0.001, ****p < 0.0001 by two-tailed, paired t-test (B, C, F and G).

Next, we transfected HEK293 cells with GFP-tagged constructs of WT and p.Y1816C mutant SORLA and quantified their surface expression levels using flow-cytometry analysis (**Fig. 2E**). First, we gated for GFP-positive cells, such that readouts were independent of any difference in applied transfection efficiency. Upon immunostaining of the transfected and non-permeabilized cells using a polyclonal antibody for the SORLA luminal domain (pAb 5387) we found a significant lower expression of p.Y1816C mutant protein compared to WT at the cell surface (median fluorescence intensity of SORLA surface positive cells relative to WT (%): 100 ± 0 (WT) vs 29.3 ± 4.8 (p.Y1816C); p = 0.0015) (**Fig. 2F**). Moreover, we found that a lower number of cells expressing the mutant protein passed the threshold value for being positive for SORLA at the cell surface (cell count of SORLA surface positive cells relative to WT (%): 100 ± 0 (WT) vs 35.1 ± 2.8 (p.Y1816C); p = 0.0006) (**Fig. 2G**). The decreased level of mutant SORLA at the cell surface provides a quantitative explanation for the decrease of the shed sSORLA fragment in the cell medium.

In conclusion, we found that the p.Y1816C mutant in HEK293 cells does not mature properly and shedding is significantly decreased to ∼30% compared to cells transfected with the WT receptor construct. Moreover, our flow cytometry measurements indicated that trafficking of p.Y1816C mutant protein to the cell surface was decreased to ∼30% compared to cells transfected with the WT receptor construct.

The observed decreases in cell surface localization and receptor maturation suggest a change in the intracellular trafficking of the mutant protein. We therefore set out to determine whether the p.Y1816C mutant receptor locates to different intracellular compartments compared to the WT receptor. We performed immunocytochemistry to stain HEK293 cells transfected with untagged WT or mutant SORLA constructs using markers of the endoplasmic reticulum (Calnexin), early endosomes (EEA-1), or retromer-coated endosomal structures (the vacuolar protein sorting 35, VPS35). We visualized the co-localization between SORLA and subcellular markers with confocal microscopy (**Fig. 3**). In line with the decreased protein maturation, we observed that, compared to WT SORLA, the co-localization of mutant SORLA with Calnexin was increased, suggesting that p.Y1816C might impair receptor folding and efficient export out of the ER (Manders coefficients: 0.35 ± 0.1 (WT) vs 0.47 ± 0.2 (p.Y1816C); p = 0.0002) (**Fig. 3A-B**). Concomitantly, we observed a reduced co-localization of the mutant protein with both EEA-1 (Manders correlation coefficients: 0.4 ± 0.15 (WT) vs 0.28 ± 0.15 (p.Y1816C); p = 0.0002) (**Fig. 3C-D**) and VPS35 (Manders correlation coefficients: 0.5 ± 0.1 (WT) vs 0.39 ± 0.15 (p.Y1816C); p = 0.0011) (**Fig. 3E-F**). Despite the increased ER localization and the decreased localization to endosomal structures, we observed only a ∼30% reduction in co-localization between the p.Y1816C mutant and VPS35 compared to the WT receptor. However, with sSORLA shedding and flow-cytometry analysis of surface expression we observed a much stronger (∼70%) decrease of the mutant receptor compared to WT receptor. This lead us to speculate that the p.Y1816C mutant is additionally impaired also in its transport out of the endosome to the cell surface.

**FIGURE 3:**
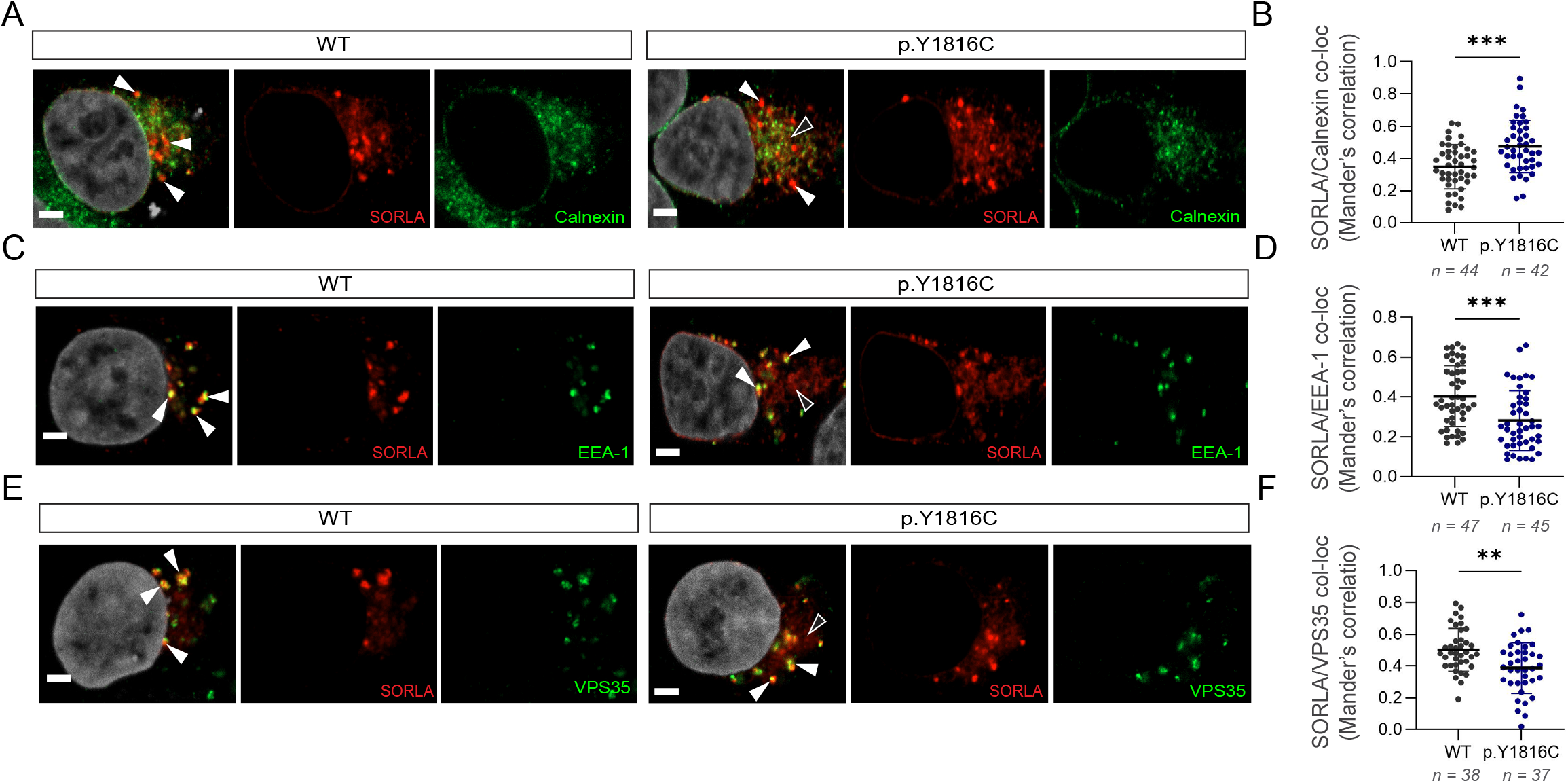
Intracellular localization of SORLA WT and p.Y1816C in HEK293 cells. Representative immunofluorescence images of HEK293 cells transfected with WT (*left*) or p.Y1816C (*right*) immunostained for SORLA (red) and Calnexin (A), EEA-1 (C) or VPS35 (E) (green) counterstained with DAPI (grey), scale bars, 2 µm. Graphs illustrate quantification of colocalization between SORLA and Calnexin (B), EEA-1 (D) or VPS35 (F) as shown by Mander’s correlation coefficients. Images were acquired from two independent experiments for each condition (Calnexin (B) (WT: n = 44; p.Y1816C: n = 42), EEA-1 (D) (WT: n = 47; p.Y1816C: n = 45) and VPS35 (F) (WT: n = 38; p.Y1816C: n = 37)). Error bars indicate mean with SD. All data were analyzed by parametric statistical tests. *p < 0.05, **p < 0.01, ***p < 0.001, ****p < 0.0001 by two-tailed, unpaired t-test (B, D and F).

We recently showed that SORLA forms dimers within endosomes, that the dimerization is in part mediated by the 3Fn-domains, and that the SORLA dimer binds the retromer complex ^22^. We therefore speculated whether the p.Y1816C mutation could impair dimerization making the mutant receptor ineffective for the retromer-dependent recycling to the cell surface.

We next generated two SORLA minireceptor constructs consisting of the six 3Fn-domains, transmembrane region and cytoplasmic tail with C-terminal Venus1 (V1) and Venus2 (V2) tags (3Fn-mini-V1 and 3Fn-mini-V2). We also introduced the p.Y1816C mutation in these constructs to study whether the 3Fn-domain mediated dimerization was abolished after introduction of the p.Y1816C variant. First we transfected HEK293 cells with either WT or p.Y1816C 3Fn-mini-Venus constructs and performed the bimolecular fluorescence complementation (BiFC) assay followed by immunocytochemistry to stain for SORLA and VPS35. Upon transfection of HEK293 cells with the WT 3Fn-mini-Venus constructs we observed a clear and focused signal of the reconstituted Venus protein that represents homodimerization (**Fig. 4A-C**). Moreover, there was a strong overlap between the WT 3Fn dimer and the VPS35 subunit of retromer marking the endosome as observed by confocal microscopy imaging (**Fig. 4B**). In contrast, there was only a neglible Venus signal after transfection with the p.Y1816C 3Fn-mini-Venus constructs indicating that homodimerization was abrogated in the mutant (**Fig 4D-F**). However, for both the WT and p.Y1816C 3Fn-mini-Venus constructs we detected a high co-localization between the SORLA staining (3Fn-mini-Venus monomers and dimers) and VPS35 (Mander’s correlation coefficients: 0.56 ± 0.13 (WT) vs 0.41 ± 0.14 (p.Y1816C); p = < 0.0001), but less overlap for the mutant compared to the WT (**Fig. 4G**). Additionally, we quantified the co-localization between the WT Venus signal and VPS35 (Mander’s correlation coefficients: 0.83 ± 0.1 (WT)) revealing a remarkably high overlap between WT 3Fn homodimers and VPS35 (**Fig. 4H**). Thus, this confirms that 3Fn-mediated dimerization occurs mostly in retromer-positive endosomal compartments and that homodimerization between SORLA’s 3Fn domains is abolished in the p.Y1816C mutated protein. Together these findings demonstrate that the 3Fn-mediated dimerization is strongly imparied by the p.Y1816C mutation.

**FIGURE 4:**
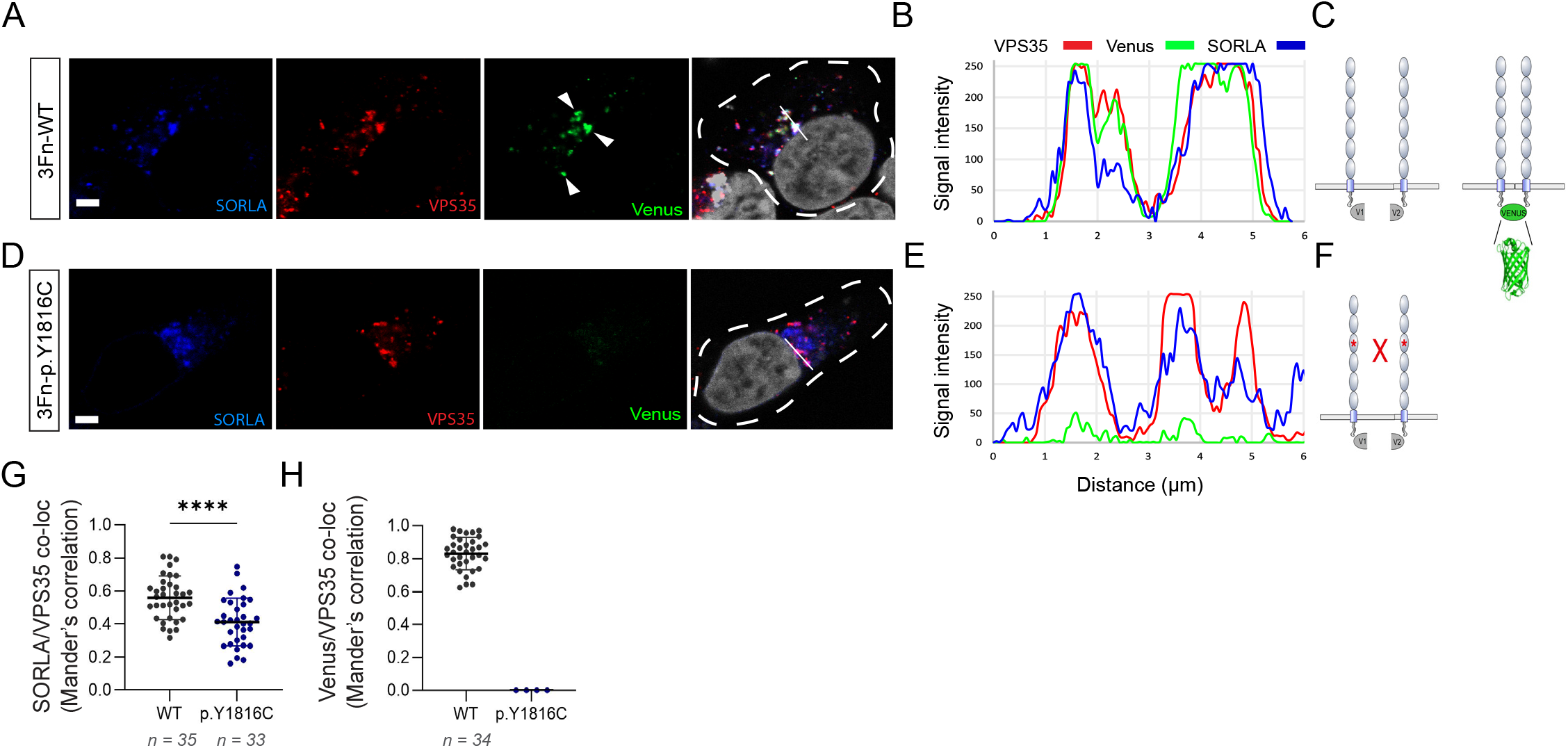
Intracellular localization of WT and p.Y1816C 3Fn homodimers in HEK293 cells. Representative immunofluorescence images of HEK293 cells transfected with 3Fn WT (A) or p.Y1816C (D) Venus-1 and Venus-2 constructs (green) co-stained with SORLA (blue), VPS35 (red) and DAPI (grey). Green venus signals represent 3Fn SORLA dimers only. Profile intensity plots showing comparison of fluorescence intensity versus distance (µm) over a fixed line spanning venus (green), VPS35 (red) and SORLA (blue) positive structures in a representative WT (B) and p.Y1816C cell (E). Schemtaic representations demonstrating reconstitution of the Venus fluorescent protein upon dimerization between WT 3Fn (C) or abrogation of the Venus signal in the p.Y1816C 3Fn mutant (F). Graphs illustrating quantifications of colocalization between VPS35 and either SORLA staining (G) or 3Fn Venus (H) as shown by Mander’s correlation coefficient. Images were acquired from two independent experiments for each condition (SORLA staining (G) (WT: n = 35; p.Y1816C: n = 34), 3Fn Venus (H) (WT: n = 34; p.Y1816C: n = 4 arbritary units)). For colocalization analysis between VPS35 and Venus signal from the p.Y1816C 3Fn mutant (G) Mander’s coeeficients were set as 0 as arbritary numbers since there was virtually no Venus signal. Scale bars, 2 µm. Error bars indicate mean with SD. All data were analyzed by parametric statistical tests. *p < 0.05, **p < 0.01, ***p < 0.001, ****p < 0.0001 by two-tailed, unpaired t-test (G).

To investigate how p.Y1816C impact dimerization of the full-length SORLA receptor, we repeated the BiFC assay using the two Venus1- and Venus2-tagged SORLA constructs that were previously described for the full-length receptor ^22^. Again, we introduced the p.Y1816C mutation into each of the two reporters. Transfection of the two Venus-tagged WT SORLA constructs into HEK293 cells, provided a clear and focused signal of the reconstituted Venus protein (**Fig. 5A**) that showed a strong overlap with VPS35 as observed by confocal microscopy imaging (**Fig. 5B**) which was also reported previously ^22^. We also observed signal for dimer formation by the p.Y1816C mutant full-length homodimer in cells transfected with Venus-tagged SORLA p.Y1816C constructs (**Fig. 5C**), but in clear contrast to the WT full-length homodimer, the signal for the mutant dimer was distributed across most of the cellular cytoplasm in a pattern compatible with ER staining and with virtually no co-localization with retromer (**Fig. 5D**). While the absence of co-localization between dimerized mutant SORLA and retromer can explain the concomitant strong decrease in cell surface shedding, we were surprised to yet observe a clear Venus signal for the mutant full-length dimer. To determine where in the cell the mutant can form a dimer, we immunostained cells transfected with the Venus-tagged WT or p.Y1816C SORLA constructs with Calnexin to visualize the ER. By confocal microscopy imaging we observed again a focused Venus signal in cells transfected with the Venus-tagged WT constructs (**Fig. 5E**), but only a small overlap between the WT dimer and Calnexin (**Fig. 5F**). However, when transfecting cells with Venus-tagged p.Y1816C constructs, we observed clear Venus signals for the p.Y1816C mutant homodimer (**Fig. 5G**) showing a high overlap with Calnexin (**Fig. 5H**).

**FIGURE 5:**
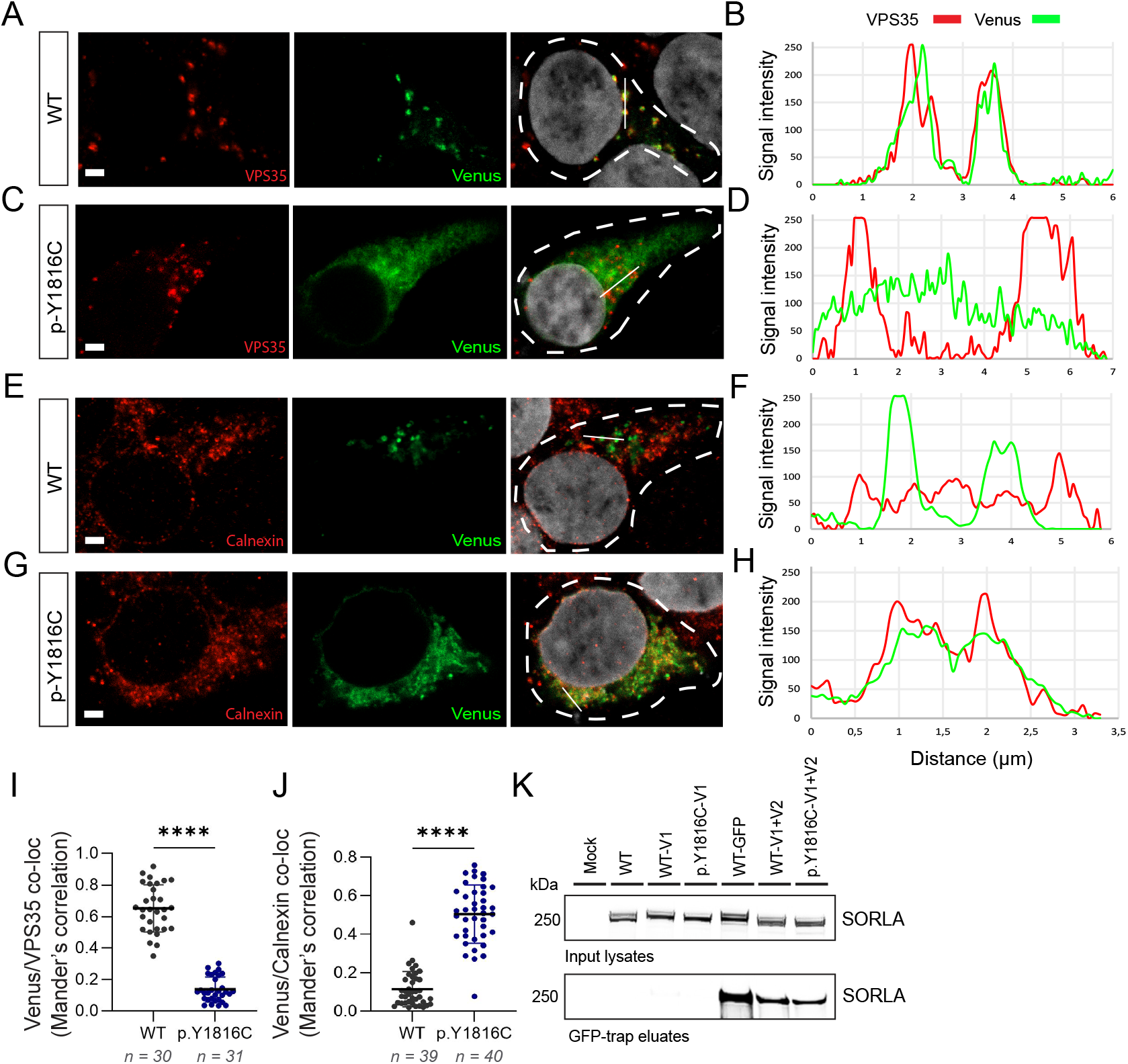
Intracellular localization of full-length SORLA WT and p.Y1816C homodimers in HEK293 cells. Representative immunofluorescence images of HEK293 cells transfected with SORLA WT (A) or p.Y1816C (C) Venus-1 and Venus-2 constructs (green) co-stained with VPS35 (red) and DAPI (grey). Green venus signals represent SORLA dimers only. Profile intensity plots showing comparison of fluorescence intensity versus distance (µm) over a fixed line spanning VPS35 (red) and venus (green) positive structures in a representative WT (B) and p.Y1816C cell (D). Representative immunofluorescence images of HEK293 cells transfected with SORLA WT (E) or p.Y1816C (G) Venus-1 and Venus-2 constructs (green) co-stained with Calnexin (red) and DAPI (grey). Green venus signals represent SORLA dimers only. Profile intensity plots showing comparison of fluorescence intensity versus distance (µm) over a fixed line spanning Calnexin (red) and venus (green) positive structures in a representative WT (F) and p.Y1816C cell (H). Graphs illustrating quantifications of colocalization between SORLA dimers (venus) and VPS35 (I) or Calnexin (J) as shown by Mander’s correlation coefficient. Images were acquired from two independent experiments for each condition (VPS35 (I) (WT: n = 30; p.Y1816C: n = 31), Calnexin (J) (WT: n = 39; p.Y1816C: n = 40)). Scale bars, 2 µm. (K) GFP-trap immunoprecipitation analysis of lysates from HEK293 cells transfected with SORLA-WT-V1, SORLA-p.Y1816C-V1, SORLA-WT-GFP, SORLA-WT-V1+V2 and SORLA-p.Y1816C-V1+V2. Input lysates and precipitated proteins were detected by WB using SORLA (LR11) antibody. Error bars indicate mean with SD. All data were analyzed by parametric statistical tests. *p < 0.05, **p < 0.01, ***p < 0.001, ****p < 0.0001 by two-tailed, unpaired t-test (I and J).

Quantification of the co-localization between the full-length WT and p.Y1816C dimers and VPS35 indicated an ∼80% difference between mutant and WT SORLA (Manders coefficients: 0.65 ± 0.15 (WT) vs 0.14 ± 0.08 (p.Y1816C); p < 0.0001) (**Fig. 5I**) suggesting that the mutation impairs the physiological relevant dimer formation by the 3Fn-domains in endosomes that allows the receptor to be recycled to the cell surface. By quantifying the co-localization between the full-length WT and p.Y1816C homodimers and Calnexin we found that the mutant homodimer is highly located to this compartment (Mander’s correlation coefficients: 0.09 ± 0.1 (WT) vs 0.53 ± 0.2 (p.Y1816C); p < 0.0001) (**Fig. 5J**). To confirm that the mutant full-length receptor forms a dimer, we confirmed efficient dimerization using the GFP-trap approach (**Fig. 5K**). These findings demonstrate that the p.Y1816C mutant dimerizes in the ER, which may explain the slight increase we observed for the co-localization of full-length mutant with Calnexin in the ER (ie. **Fig. 3A**). We speculate that dimerization of the mutant in the ER is mediated by its VPS10p-domains.

Previous studies indicated that *SORL1* is haploinsufficient ^7^ and that a 50% reduction in SORLA expression, due to a truncating *SORL1* variant, leads to endosome swelling in iPSC-derived neurons ^10, 11^, which was confirmed, *in vivo* in neurons from *SORL1*^+/-^ minipigs (i.e. heterozygous *SORL1* k.o) ^12^. More recently, also the missense variants p.E270K, p.Y141C, and p.G511R in iPSC models have been suggested to cause endosome enlargements, although the exact mechanism remains to be determined ^35^.

Therefore, we introduced the p.Y1816C variant into an iPSC model system, and determined whether this leads to the cardinal cellular phenotype of impaired SORLA activity. We generated SORLA isogenic allelic series from a parental iPSC line (schematized in **Fig. 6A**). We modified a well-established i3N iPSC line with doxycycline-inducible NGN2 expression using a CRISPR/Cas9 gene-mediated strategy. As described previously, i3N iPSCs can be differentiated into mature neurons in approximately 17 days ^36^. Upon generation of SORLA knock out (KO) and SORLA knock in for p.Y1816C (p.Y1816C), we first verified the SORLA expression in selected clones (**Fig. 6B**). The level of SORLA in p.Y1816C iPSCs was similar to the control WT iPSCs, while the level of SORLA in the KO iPSCs was undetectable (**Fig. 6B-C**). Subsequently, we induced i3N iPSCs to differentiate into neurons for 17 days to determine the endosome size using immunocytochemistry. We detected a significantly elevated percentage of EEA1-positive enlarged structures in both p.Y1816C and KO neurons compared to control WT neurons (**Fig. 6D**). Specifically, the percentage of EEA1-positive early endosomes larger than 0.5µm^2^ was 16% ± 3.0 in p.Y1816C, 20% ± 1.7 in KO, and 7.4% ± 0.98 in WT neurons (**Fig. 6E**). This elevated percentage of enlarged early endosomes was also reflected by increased levels of EEA-1 protein as detected by WB (**Fig. 6F-G**). Our data thus confirm that the SORLA p.Y1816C variant, similarly to other pathogenic loss-of-function (LOF) variants and SORLA-KO, leads to endosome swelling in iPSC-derived human neurons.

**FIGURE 6:**
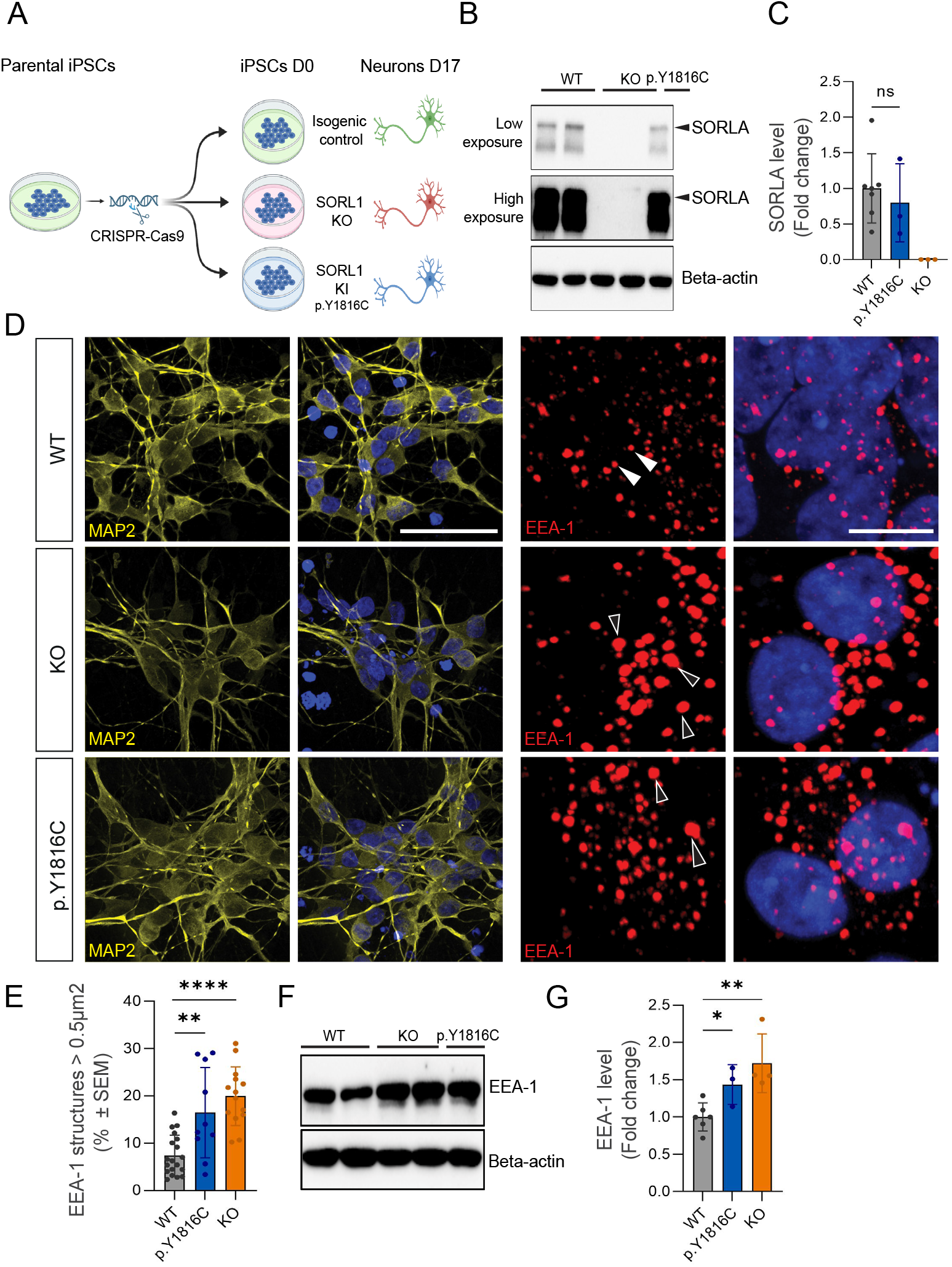
Endosome enlargement in iPSC-derived neurons with *hemizygous* SORLA p.Y1816C mutation. (A) Schematic representation of the generation of isogenic allelic series of iPSCs with hemizygous SORLA p.Y1816C mutation and SORLA KO. (B) Representative WB of SORLA protein levels in iPSCs and *(C)* bar graph showing quantification of at least three independent WB replicates. Quantification includes two isogenic clones of SORLA-WT iPSCs, two isogenic clones of SORLA-KO iPSCs and one clone of hemizygous SORLA p.Y1816C (KI) iPSCs. *(D)* Representative immunofluorescence images of WT (*top*), SORLA KO (*middle*), and SORLA KI p.Y1816C (*bottom*) neurons stained with MAP2 (yellow). Endosomes were visualized by EEA-1 (red). Nuclei were counterstained with DAPI (blue). *(E)* Quantification of immunofluorescence images demonstrates increased EEA-1 volume in SORLA KO and SORLA KI p.Y1816C neurons compared with WT. n = 3 images (50-70 cells). *(F)* Representative WB and *(G)* quantification of the protein level of EEA-1 in the SORLA WT, SORLA KO, and SORLA KI p.Y1816C iPSCs. Quantification includes two isogenic clones of WT (n=5), two isogenic clones of KO genotype (n=3) and one isogenic clone for KI genotype (n = 2). All values represent mean ± SD. All normally distributed data were analyzed using two-tailed unpaired t tests. *p < 0.05, **p < 0.01, ***p < 0.001, and ****p < 0.0001 (C, E and G).

## DISCUSSION

With this work, we provide genetic, biochemical and cell biological evidence that the rs772677709 variant in *SORL1*, which leads to a p.Y1816C substitution, affects SORLA receptor activity and should be considered pathogenic. The segregation of the rs772677709 variant with early-onset AD in three unrelated families, suggests that this *SORL1* variant leads to autosomal dominant AD. Besides the three described pedigrees, a recent large exome sequencing case-control study identified this mutation in an additional five unrelated AD cases, most with an early-onset of AD ^8^. The p.Y1816C substitution affects a highly conserved residue located within the 3rd 3Fn-domain, which is part of a structurally important motif known as the ‘*tyrosine corner*’ ^33^. It has previously been identified that the ‘*tyrosine corner*’ is not a direct folding nucleus, but rather this structure adds to the stability of the folded domain ^37^. Here, we demonstrated that maturation of the mutant receptor is impaired, expression at the cell surface and shedding of the soluble sSORLA fragment is decreased, and that endosomes are enlarged in neurons with the p.Y1816C mutation. These are all parameters that depend on SORLA’s ability to traffic to the endosome and subsequently to the cell surface ^18^, and which so far seem unifying defects for pathogenic *SORL1* variants ^19, 20, 34^.

We recently showed that WT SORLA can dimerize at two different domains: the 3Fn-domain and the VPS10-domain ^22^. However, these dimers (or possibly multimers) are observed only when SORLA is in the endosome, bound to VPS26 of the retromer complex. In fact, we did not observe dimers or multimers of WT SORLA in the ER, likely because only a negligible fraction of WT SORLA is located in this compartment. However, another possibility is that WT SORLA adopts a conformation that is unable to form dimers when located in the ER as well as during transport from ER to Golgi to Endosomes. Entering the endosome may induce a conformational change allowing SORLA to dimerize by its 3Fn-domains and to interact with the (dimeric) retromer cargo-recognition complex (i.e. VPS26) and to engage in trafficking of endosome cargo.

It is notable that homologous pathogenic variants exist in other proteins with 3Fn-domains, in which a conserved tyrosine is replaced with a cysteine in the *tyrosine corner* ^34^: p.Y784C in L1CAM, p.Y973C in Fibronectin, p.Y226C in the Growth Hormone receptor, p.Y818C in the Insulin receptor, and p.Y611C in the Angiopoietin receptor Tie2. Notably, these proteins are all active in their dimer conformation and, in parallel with the observed effects of the SORLA p.Y1816C mutant, the tyrosine substitutions do not lead to dramatic changes in their subcellular localization, but rather abolish their biological function relying on dimerization. Functional studies of the mutant Tie2 protein, which causes primary congenital glaucoma (OMIM: GLC3E) ^38^, indicated that while the Y611C substitution did not lead to decreased cell surface expression compared to WT Tie2, the mutant was unable to respond to ligand-mediated activation likely due to impaired dimer and multimer formation ^38^. This strongly parallels our observations of the SORLA p.Y1816C mutant for which we observe only a modest decrease in the endosome localization, but for which the overlap between the SORLA-Venus dimer with retromer is almost completely lost. This suggests that similar to Tie2, SORLA cannot perform its physiological function (ie. dimerization and retromer-dependent endosome recycling of cargo) when having the tyrosine corner mutated due to instability of the domain. Taken together, homologous substitutions in protein homologs not only provide genetic support that the p.Y1816C variant in *SORL1* is pathogenic, but also provides independent supporting evidence for the observed mechanism of pathogenicity.

*APOE* genotype strongly affects the age at onset of AD ^39^, and we and others have previously shown that carrying an *APOE*-e4 allele, expedites the age at onset of *SORL1* variant carriers ^40, 41^. The probands had onset ages ranging from 50 – 69 years, and all three carried the *APOE*-e3/e4 genotype which likely expedited the age at onset in these individuals relative to non-*APOE*-e4 carriers. Furthermore, age at onset of sporadic AD and the penetrance of familial AD is also affected by other genetic variants ^39, 42–45^, which, in addition to possible differences in lifestyle, might explain that the sibling of the index patient from family 1, who carried the neutral *APOE* e3/e3 genotype was not affected with AD when last seen at an age between 55 and 59 years.

In conclusion, the rs772677709 variant in *SORL1*, which leads to a p.Y1816C substitution represents one of two autosomal dominant variants identified thus far in the *SORL1* gene, the other being the p.D1545V Icelandic mutation ^13^. The function of SORLA in carriers of the p.Y1816C mutant is differentially impaired compared to carriers of the p.D1545V mutant. The latter affects the ‘*calcium cage*’ of CR-domains, which leads to a misfolded protein, causing retention in the early secretory compartments ^13, 46^. In contrast, the mutated ‘*tyrosine corner’* in the p.Y1816C mutant still allows proper folding of the 3Fn-domain, but renders it in an unstable conformation. Only a minor fraction of the mutated protein is retained in the ER, while the majority of the protein mutant still sorts relatively efficiently to its correct cellular destination. However, once there, failure to dimerize and bind the retromer at the endosome impairs SORLA function. In summary, our work provides additional evidence that certain missense variants in *SORL1* impair SORLA function such that this leads to autosomal dominant AD, and that *SORL1* should be regarded a fourth autosomal dominant gene for Alzheimer’s disease.

## Data Availability

All data produced in the present study are available upon reasonable request to the authors

## Acknowledgement

We thank Sandra Bonnesen for excellent technical assistance and Ceren Akcasoy for skillful assistance with site-directed mutagenesis. We also thank Jan Asad for help in setting up the flow cytometry experiments.

Flow cytometry was performed at the FACS Core Facility, Aarhus University, Denmark and we thank the facility staff for excellent assistance with the flow cytometry experiments.

The authors also acknowledge AU Health Bioimaging Core Facility and CELLIM Cellular Imaging Core Facility CEITEC supported by the MEYS CR (LM2018129 Czech-BioImaging) for the use of equipment and support of the imaging facility.

H.H., O.M.A., and D.B. are a part of the EU Joint Programme-Neurodegenerative Disease Research (JPND) Working Group SORLA-FIX under the 2019 ‘‘Personalized Medicine’’ call (JPND2019-466-197, ZonMW 733051110, Danish Innovation Foundation and the Velux Foundation Denmark, and the Ministry of Education, Youth and Sports of the Czech Republic no. 8F20009). H.H., is a recipient of ABOARD, a public-private partnership receiving funding from ZonMW (#73305095007) and Health∼Holland, Topsector Life Sciences & Health (PPP-allowance; #LSHM20106). H.H. was supported by the Hans und Ilse Breuer Stiftung (2020) and the HorstingStuit Foundation (2018). O.M.A is supported by Novo Nordisk Foundation (#NNF20OC0064162), the Alzheimer’s Association (ADSF-21-831378-C), the Independent Research Fund Denmark (#3101-00065B) and the Danish Alzheimer’s Research Foundation (recipient of the 2022 Basic Research Science Award). Research of K.S. is supported in part by the SAO and UA special research fund. SvdL received funding from ZonMW (#733050512). J.R. is supported by the Czech Health Research Council (AZV project No. NU22J-08-00075). D.B. is suported by project nr. LX22NPO5107 (MEYS): Financed by European Union – Next Generation EU. L.B. and R.G. were supported by the Italian Ministry of Health (Ricerca Corrente).

## Notes

### Competing Interest Statement

O.M.A. is a consultant for Retromer Therapeutics and has equity.

### Author Declarations

The Medical Ethics Committee of the Amsterdam University Medical Center gave ethical approval for this work.

