## Supplementary Information for "The *SORL1* p.Y1816C variant causes impaired endosomal dimerization and autosomal dominant Alzheimer’s disease"

### Jensen
### Supplementary Table S1

#### i3N induction medium (IM)

| ITEM | COMPANY | IDENTIFIER | STORAGE CONCENTRATION | FINAL CONCENTRATION |
|---|---|---|---|---|
| DMEM/F12 | Thermo Fisher Scientific | Cat# 21331046 | basal medium | basal medium |
| HEPES | Thermo Fisher Scientific | Cat# 15630106 | 1M | 15mM |
| N-2 Supplement | Thermo Fisher Scientific | Cat# 17502048 | 100X | 1X |
| GlutaMAX™ | Thermo Fisher Scientific | Cat# 35050061 | 100X | 1X |
| MEM Non-Essential Amino Acids | Thermo Fisher Scientific | Cat# 11140035 | 100X | 1X |

#### i3N cortical neuron medium (CNM)

| ITEM | COMPANY | IDENTIFIER | STORAGE CONCENTRATION | FINAL CONCENTRATION |
|---|---|---|---|---|
| BrainPhys™ Neuronal Medium | STEMCELL Technologies | Cat# 5790 | basal medium | basal medium |
| B-27™ Supplement, minus vitamin A | Thermo Fisher Scientific | Cat# 12587-010 | 50X | 1X |
| BDNF | PeproTech | Cat# 450-02 | 10 µg/mL | 10 ng/mL |
| NT3 | PeproTech | Cat# 450-03 | 10 µg/mL | 10 ng/mL |
| Laminin Mouse Protein | Thermo Fisher Scientific | Cat# 23017015 | 1 mg/mL | 1 ug/mL |

#### qPCR primers

| GENE | NCBI ID | FORWARD SEQUENCE 5’-3’ | REVERSE SEQUENCE 5’-3’ |
|---|---|---|---|
| *GAPDH* | 2597 | AGCCACATCGCTCAGACAC | GCCCAATACGACCAAATCC |
| *SORL1* | 6653 | CCAACCTGAAGATGGGTCATA | ACAGCAGCAACATCCGTAGAT |

#### WB antibodies

| ANTIBODY | DILUTION | COMPANY | IDENTIFIER |
|---|---|---|---|
| β-Actin (8H10D10) | 1:10,000 | Cell Signaling Technology | Cat# 3700; RRID: AB_2242334 |
| LR11 (SORLA) | 1:4,000 | BD Biosciences | Cat# 611861; RRID: AB_399341 |
| EEA1 (G-4) | 1:500 | Santa Cruz Biotechnology | Cat# sc-137130; RRID: AB_2246349 |
| Anti-mouse IgG, HRP | 1:3,000 | Cell Signaling Technology | Cat# 7076; RRID: AB_330924 |

**Jensen**
**Supplementary Table S2**

| | | Genomic location | | | | Family 3 proband | | Family 1 proband | | Family 2 proband | | CON | | CON | | CON | |
|---|---|---|---|---|---|---|---|---|---|---|---|---|---|---|---|---|---|
| **ID** | **Marker** | **Start** | **End** | **Distance to SORL1 (kb)** | **Heterozygosity** | | | | | | | | | | | | |
| **1** | D11S4127 | 117.644.118 | 117.644.500 | 3679 | 0,72 | 94 | 102 | 100 | 100 | 96 | 98 | 96 | 100 | 92 | 100 | 96 | 98 |
| **2** | D11S924 | 119.437.791 | 119.438.160 | 1885 | 0,70 | 248 | 250 | 250 | 250 | 246 | 246 | 246 | 246 | 250 | 254 | 248 | 250 |
| **3** | D11S925 | 120.828.211 | 120.828.546 | 495 | 0,85 | 174 | 199 | 174 | 197 | 174 | 195 | 174 | 174 | 174 | 193 | 174 | 193 |
| | SORL1 | 121.322.912 | 121.504.471 | | | SORL1 | | | | | | | | | | | |
| **4** | D11S1377 | 122.451.716 | 122.451.852 | 829 | 0,78 | 229 | 235 | 223 | 231 | 231 | 231 | 233 | 237 | 229 | 231 | 229 | 231 |
| **5** | D11S933 | 124.671.980 | 124.672.327 | 3349 | 0,80 | 413 | 423 | 417 | 421 | 413 | 421 | 413 | 427 | 421 | 423 | 423 | 423 |
| **6** | D11S975 | 125.828.395 | 125.828.653 | 4505 | 0,79 | 223 | 225 | 223 | 233 | 223 | 233 | 223 | 223 | 233 | 237 | 223 | 233 |
| **7** | D11S4110 | 126.971.669 | 126.971.788 | 5649 | 0,71 | 343 | 343 | 343 | 353 | 343 | 343 | 349 | 353 | 343 | 351 | 351 | 353 |

**Jensen**
**Supplementary Figure S1**

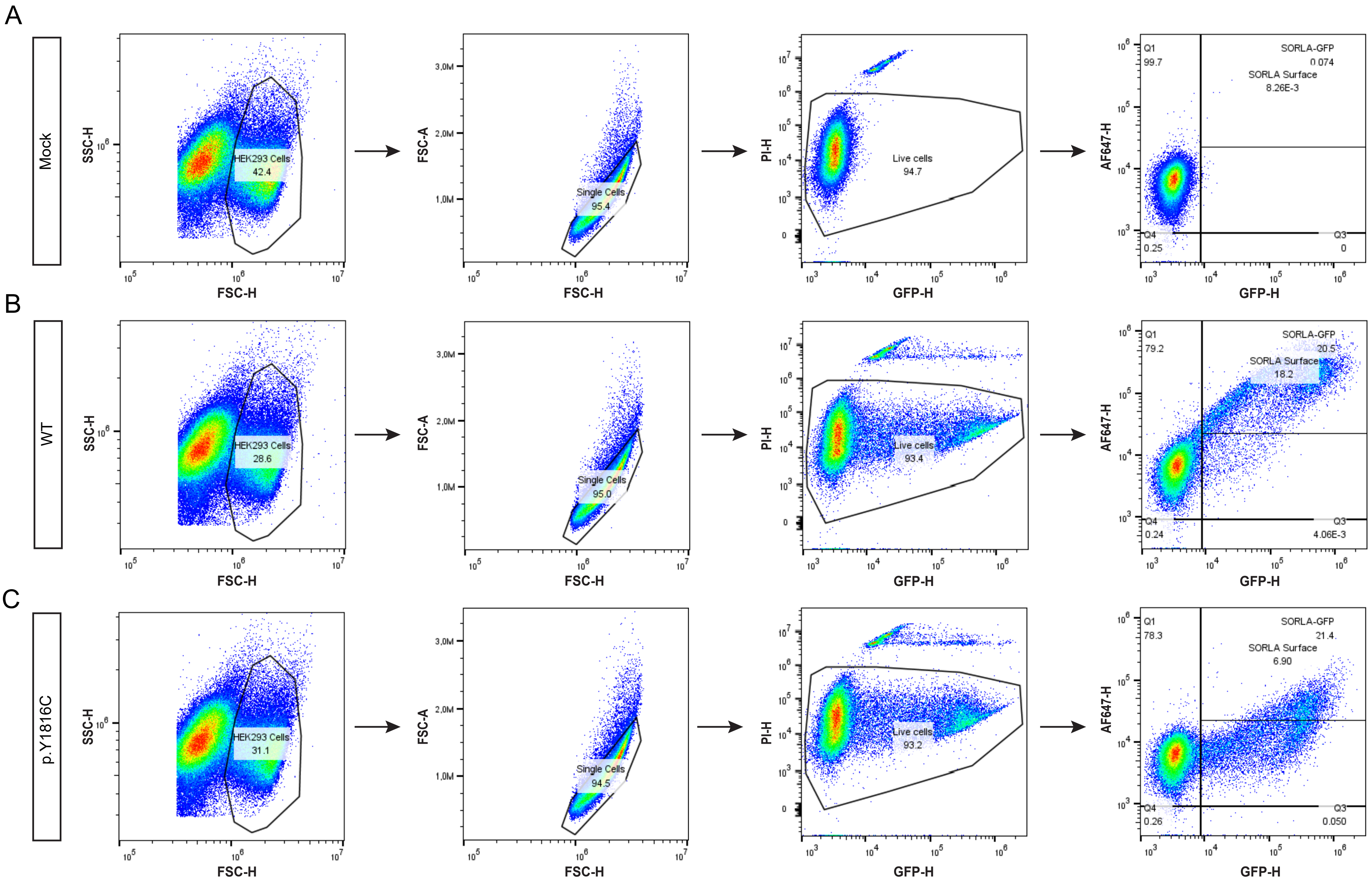

**Jensen**
**Supplementary Figure S2**

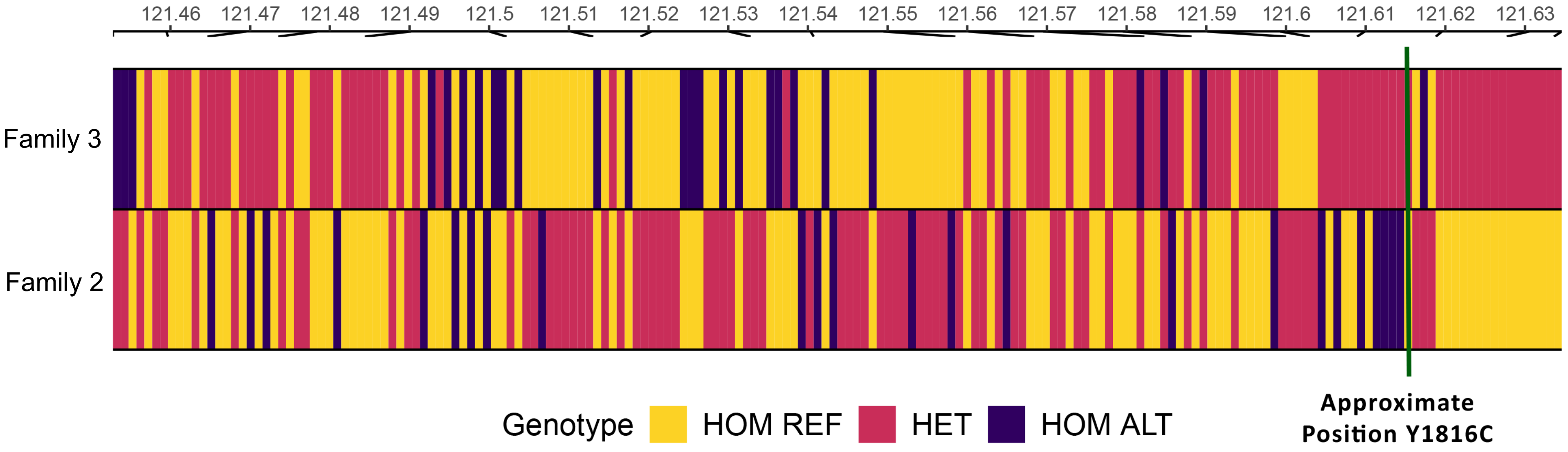
